# Sales of over the counter (OTC) codeine-containing products in the United Kingdom: a retrospective observational study

**DOI:** 10.1101/2023.11.23.23298967

**Authors:** Georgia C. Richards, Isabella Martus, Jeffrey K. Aronson, Carl Heneghan

**Affiliations:** Centre for Evidence-Based Medicine, Nuffield Department of Primary Care Health Sciences, University of Oxford, Radcliffe Observatory Quarter, Woodstock Road, Oxford, OX2 6GG, UK; Oxford Medical School, Medical Sciences Divisional Office, University of Oxford, Oxford, OX3 9DU, UK

**Keywords:** codeine, opioids, over the counter, OTC, pharmacy, dependence, misuse, abuse

## Abstract

**Background:** Codeine is a widely available opioid medicine that is on the World Health Organisation’s list of essential medicines. However, widespread access to codeine has led to its misuse, abuse, dependence, and deaths. Some countries, including France and Australia, have successfully reclassified codeine to prescription only, which is encouraging other regulators, including the UK, to reconsider the status of codeine. However, little is known about how much codeine is sold in the UK.

**Aim:** To assess national trends in sales and expenditure of codeine-containing products sold over the counter (OTC) between 2013 and 2019.

**Methods:** We conducted a retrospective observational study using electronic point-of-sales data from the human data science company *IQVIA* and population statistics from the UK’s Office of National Statistics (ONS). Descriptive statistics were used to examine the quantity, trends over time, and types of OTC codeine-containing products sold.

**Results:** 4.75 billion dosage units of codeine were sold OTC in the UK between April 2013 and March 2019, an average of 72 dosage units per UK resident. Over time, sales of codeine fell by 8%, from 12.54 dosage units per resident in 2013 to 11.48 dosage units per resident in 2019. Codeine was often sold in combination with other analgesics, amounting to 1711 tonnes of paracetamol and 96 tonnes of ibuprofen. The public spent £638 million on OTC codeine-containing products, which increased by 12% over the study period. There were 83 different types of codeine-containing products sold.

**Conclusion:** Large volumes of codeine-containing products were purchased OTC in the UK in 2013-19. To improve the safety of opioids, OTC codeine sales data should be made accessible for public health surveillance. The trends presented in this study should inform policy for the future status of codeine availability in the UK.

**Principal Investigator statement:** The authors confirm that the Principal Investigator for this paper is Dr Georgia Richards and that they had direct responsibility for the conduct of this study.

## INTRODUCTION

Codeine phosphate is one of the most widely accessible and commonly used opioids globally. In UK primary care, codeine was the most frequently prescribed opioid, and prescriptions which increased substantially between 2005 and 2017 [1]. In many countries, including the UK, codeine-containing products (e.g. co-codamol) can be purchased over the counter (OTC) without a prescription or consultation with a doctor [2].

Systematic reviews on the effectiveness of codeine report benefits for mild pain, acute postoperative pain in adults, and cough in people with cancers [3–6]. Although codeine is considered a weak opioid, it can cause adverse effects, including cardiac arrhythmias, confusion, constipation, dizziness, drowsiness, and abdominal cramps [7]. Codeine can also be abused and misused and can cause dependence and addiction [8–10]. Because of its accessibility, codeine has often been described as a gateway drug to more potent opioids, including oxycodone, morphine, heroin, and fentanyl [8]. In England and Wales, the number of deaths attributed to codeine-containing products increased from 26 deaths in 2001 to 88 deaths in 2011 and more than doubled in 2021 to 200 deaths [11].

Regulation of codeine-containing products varies globally. Most (56%; 15 of 27) of the member states in the European Union do not allow OTC sale of codeine [12]. In 2017, France reclassified codeine from OTC to prescription-only [13,14], whereafter, more falsified codeine prescriptions for cough were presented to pharmacies [15]. A questionnaire sent to 33 pharmacies in Normandy, France, showed that sales of OTC codeine products, such as *sirop Euphon®,* more than doubled between May and June 2017, the month before the restrictions were implemented [16]. Australia similarly reclassified codeine to prescription-only in 2018, which reduced codeine-related poisonings, with no change in calls to poison centers or sales for high-strength codeine [17].

In the UK, the Government is examining the status of codeine after several changes to regulations in the past two decades [18]. In 2005, pack sizes of codeine sold OTC were reduced from 100 tablets to 32 tablets and warnings were recommended to be added to patient information leaflets about addiction and the duration of use to be a maximum of three days [19].

However, an investigation in 2009 found that generic brands contained more than 32 tablets and recommendations were not systematically applied to effervescent tablets containing codeine [19]. In 2009, the Medicines and Healthcare Regulatory and Products Agency (MHRA) removed colds, influenza, coughs and sore throats as indications for codeine. It reinforced that warnings regarding addiction and the duration of three days of use must be applied to patient information leaflets for all products containing codeine, including effervescent tablets [19]. In 2013, the use of codeine was restricted in children (<12 years) and contraindicated in all children and adolescents (<18 years) who underwent a tonsillectomy, adenoidectomy, both, and any other procedure for obstructive sleep apnoea [20]. In 2020, the MHRA launched the Opioid Expert Working Group to discuss risks associated with prescribed and OTC opioids [21]. In July 2023, the MHRA opened a public consultation on the proposal to make codeine linctus, a cough syrup sold OTC, a prescription-only medicine [18].

Previous research has compared the sales of OTC codeine-containing products in 31 countries, which showed that the UK was the fourth highest-ranking [22]. However, a detailed analysis of the use of codeine OTC in the UK has not yet been published. In this study, we aimed to fill this gap by assessing the trends in sales and expenditure of OTC codeine-containing products in the UK between April 2013 and March 2019.

## METHODS

### Study Design

We designed a retrospective observational study and pre-registered the study protocol on an open repository [23].

### Data Sources

Data on codeine sold OTC are not publicly available. We obtained consumer health sales data from the human data science company, *IQVIA,* which provided quarterly sales from 1 April 2013 to March 2019. These data have been used in other published observational studies [24–27]. The data from *IQVIA* included products from the adult pain relief or cough product categories. They included the number of packs and bottles sold, the number of tablets or millilitres of liquids sold, and the dosage units. Data were collected using scan track barcodes from electronic point of sale (EPoS) store data in the UK. We obtained population data for each calendar year from the Office of National Statistics (ONS).

### Data analysis

The total volume of dosage units of codeine purchased and total public expenditure (GBP, £) for the six-year study period were calculated by combining quarterly data to estimate total dosage units sold. *IQVIA*’s “standard unit” variable was used to calculate “dosage units” of codeine to standardise the volumes of liquid and solid dosage forms. For liquids, one unit was 12 mL of codeine, and for solids, this was dependent on the dosage of codeine. *IQVIA’s* “price to public” variable was used for public expenditure, which is defined as the “pharmacy selling price or consumer purchase price or price to the public”.

Dosage units and public expenditure were divided by the UK population for each calendar year to calculate annual rates. We assessed trends over time and calculated the percentage changes. Descriptive statistics on the type of product sold, OTC formulations, pack sizes, and dose or strength of products were then calculated.

### Software and data sharing

We used Stata v16 and Python v3 in Jupyter Notebooks with pandas, seaborn, and matplotlib libraries for analysis and figures. The data provided by *IQVIA* is considered commercial and cannot be openly shared owing to our contractual agreement. We pre-registered, published, and shared our study protocol and materials on the Open Science Framework (OSF) [23,28] and our statistical code on GitHub [29].

## RESULTS

A total of 4.75 billion dosage units of codeine were sold OTC in the UK between April 2013 and March 2019 (72 dosage units per UK resident). Sales of OTC codeine fell by 8%, from 12.5 dosage units per resident in April 2013 to 11.5 units per resident in March 2019 (Figure 1).

**Figure 1.**
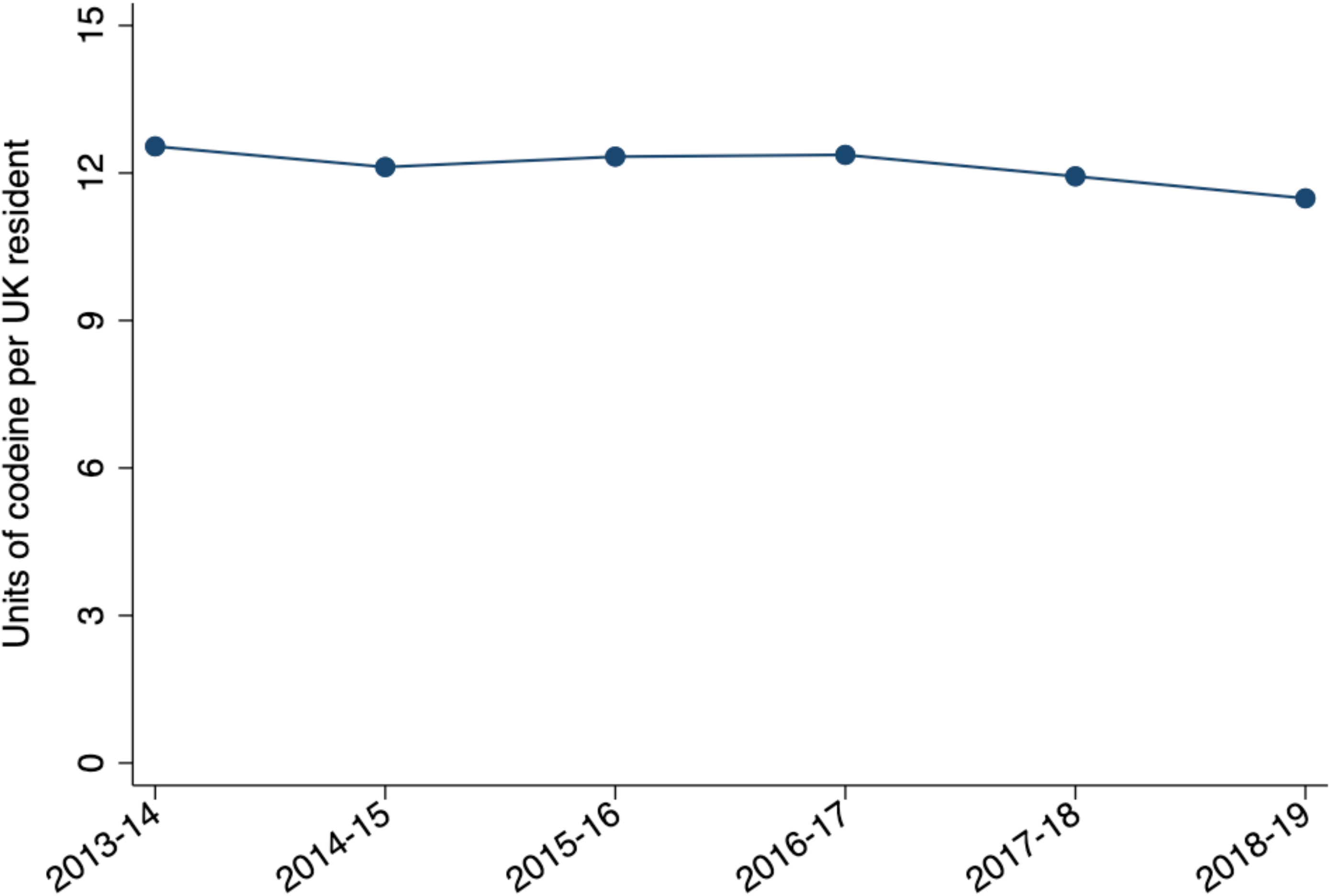
Sales of OTC codeine-containing products sold in the UK between April 2013-March 2014 and April 2018-March 2019.

There were 83 products containing codeine sold in the UK between April 2013 and March 2019. The UK sold codeine OTC in five formulations, tablets (49%), effervescent tablets (21%), and liquids (13%) being the most common (Table 1). Products contained a median of 8 mg of codeine (IQR: 8-12.8 mg), and combinations contained a median of two substances (IQR: 2-3), most (74%) containing paracetamol (Table 1). Packs had a median of 30 dosage units (IQR: 18-31 dosage units), and liquids had a median volume of 200 mL (IQR: 150-2000 mL).

**Table 1:**
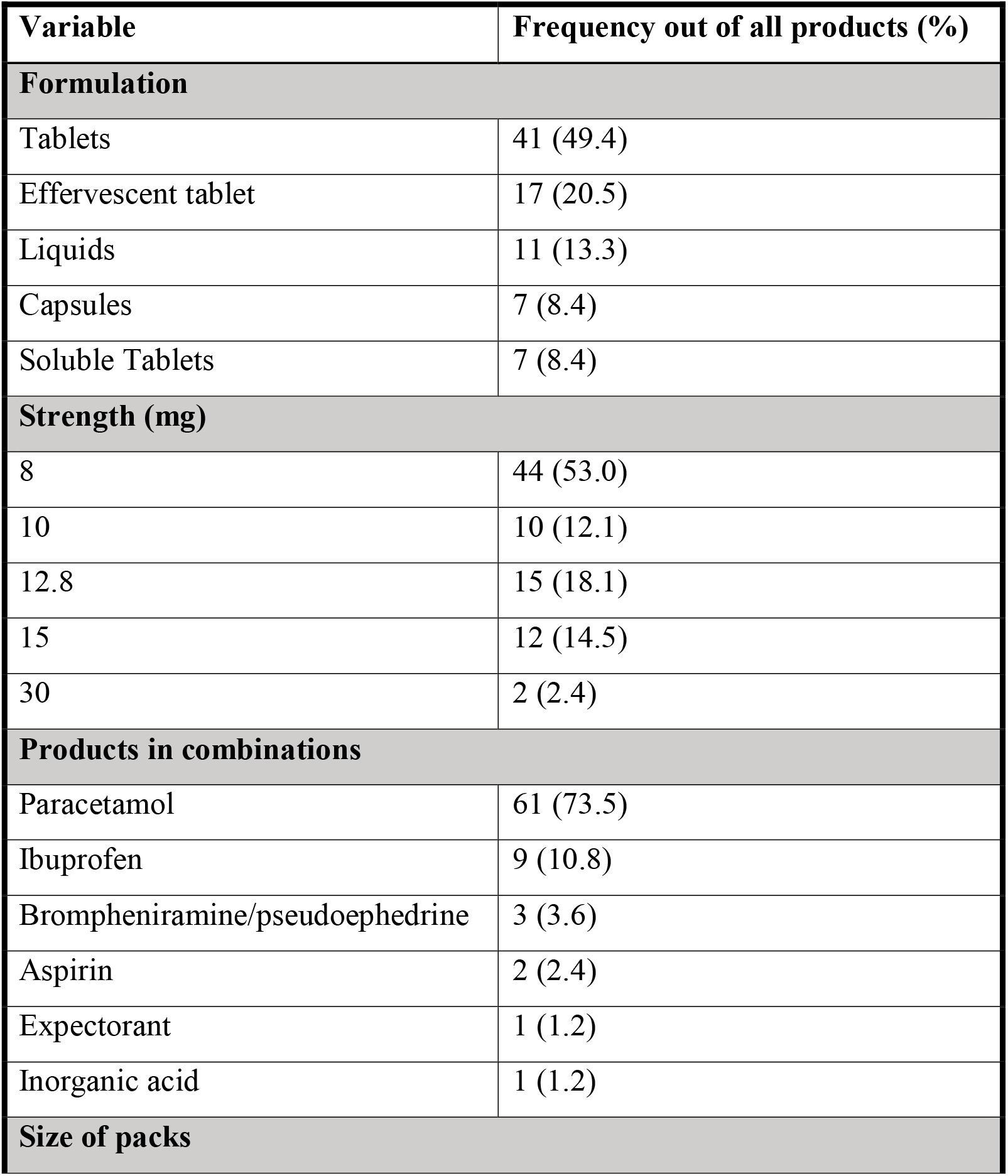
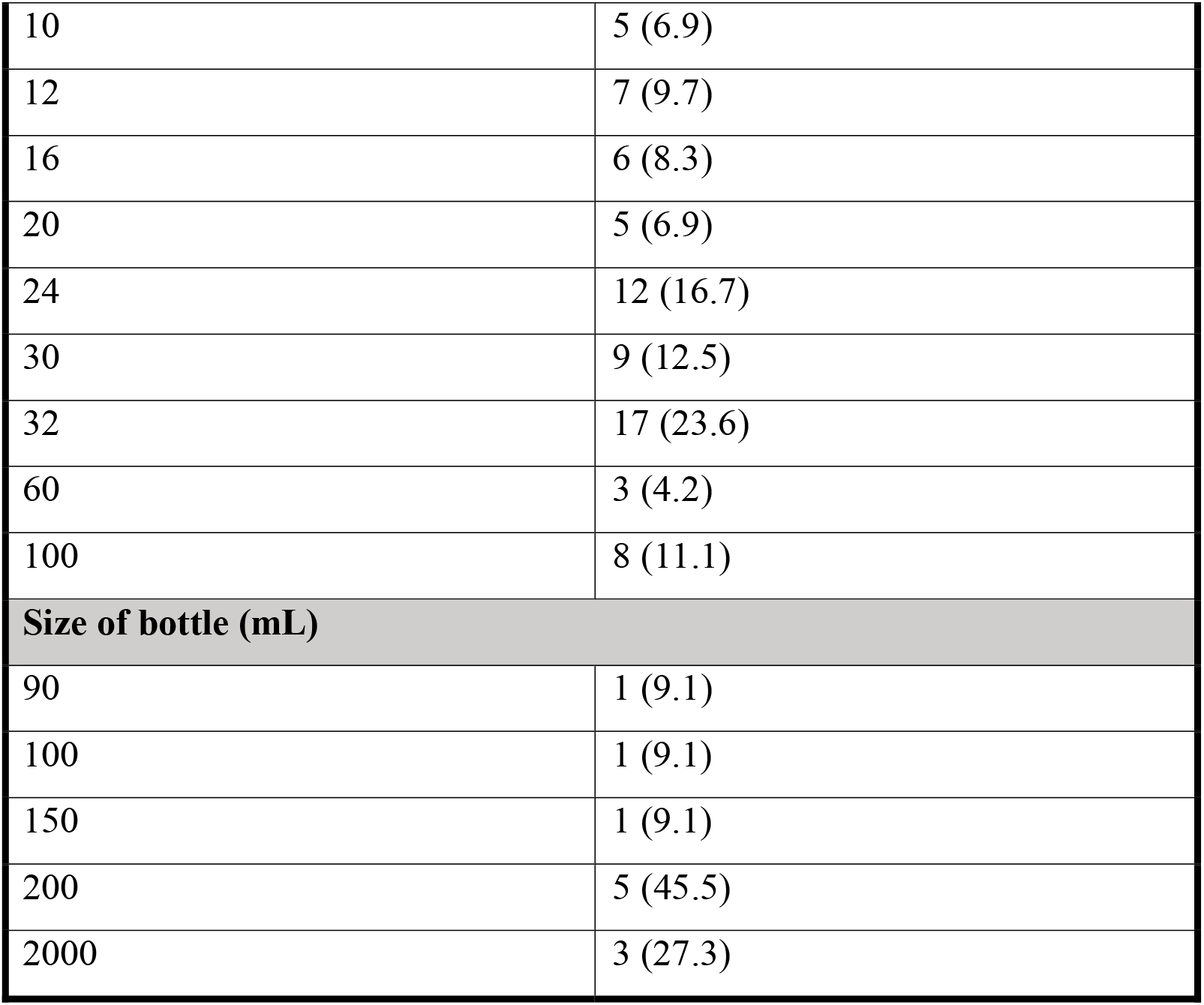
The types of products, formulations, pack sizes, and strengths of OTC codeine-containing products sold in the UK between April 2013 and March 2019.

| Variable | Frequency out of all products (%) |
| --- | --- |
| **Formulation** |  |
| Tablets | 41 (49.4) |
| Effervescent tablet | 17 (20.5) |
| Liquids | 11 (13.3) |
| Capsules | 7 (8.4) |
| Soluble Tablets | 7 (8.4) |
| **Strength (mg)** |  |
| 8 | 44 (53.0) |
| 10 | 10 (12.1) |
| 12.8 | 15 (18.1) |
| 15 | 12 (14.5) |
| 30 | 2 (2.4) |
| **Products in combinations** |  |
| Paracetamol | 61 (73.5) |
| Ibuprofen | 9 (10.8) |
| Brompheniramine/pseudoephedrine | 3 (3.6) |
| Aspirin | 2 (2.4) |
| Expectorant | 1 (1.2) |
| Inorganic acid | 1 (1.2) |
| **Size of packs** |  |
| 10 | 5 (6.9) |
| 12 | 7 (9.7) |
| 16 | 6 (8.3) |
| 20 | 5 (6.9) |
| 24 | 12 (16.7) |
| 30 | 9 (12.5) |
| 32 | 17 (23.6) |
| 60 | 3 (4.2) |
| 100 | 8 (11.1) |
| **Size of bottle (mL)** |  |
| 90 | 1 (9.1) |
| 100 | 1 (9.1) |
| 150 | 1 (9.1) |
| 200 | 5 (45.5) |
| 2000 | 3 (27.3) |

Over the 6-year study period, 46 tonnes of codeine (equivalent to 4.6 tonnes of morphine) and 10,046 kilolitres of codeine were sold OTC in the UK (Table 2). After adjusting for population, 694 mg of codeine and 151 mL of codeine were purchased for every UK resident. During this period, 1711 tonnes of paracetamol (26 g per person) and 96 tonnes of ibuprofen (1.4 g per person) were sold in combination with OTC codeine in the UK (Table 2).

**Table 2.** The volume and rate of OTC codeine-containing products and their combination products sold in the UK over six years (April 2013-March 2014 to April 2018-March 2019) by solid and liquid formulations. *data from 2019 represent sales from April 2018 to March 2019.

|  | 6-year total | 2019* total |
| --- | --- | --- |
| **Solids** |  |  |
| Volume codeine (kg) | 46108 | 7970 |
| Rate of codeine (mg/person) | 694 | 120 |
| Number of packs | 108 million | 17.5 million |
| Packs/person | 1.63 | 0.26 |
| Volume of paracetamol (kg) | 1710956 | 280810 |
| Rate of paracetamol (mg/person) | 25739 | 4230 |
| Volume of ibuprofen (kg) | 96020 | 18388 |
| Rate of ibuprofen (mg/person) | 1445 | 277 |
| **Liquids** |  |  |
| Volume of codeine (kL) | 10046 | 1291 |
| Rate of codeine mL/ person | 151 | 19 |

The public spent £638 million (£9.60 per person) on OTC codeine-containing products, increasing by 12% from £1.47 per person in April 2013 to £1.64 in March 2019 (Figure 2). One dosage unit of codeine costs an average of £0.13 per dose, which increased by 22% from £0.12 per dosage unit of codeine in April 2013 to £0.14 per dosage unit of codeine in March 2019.

**Figure 2.**
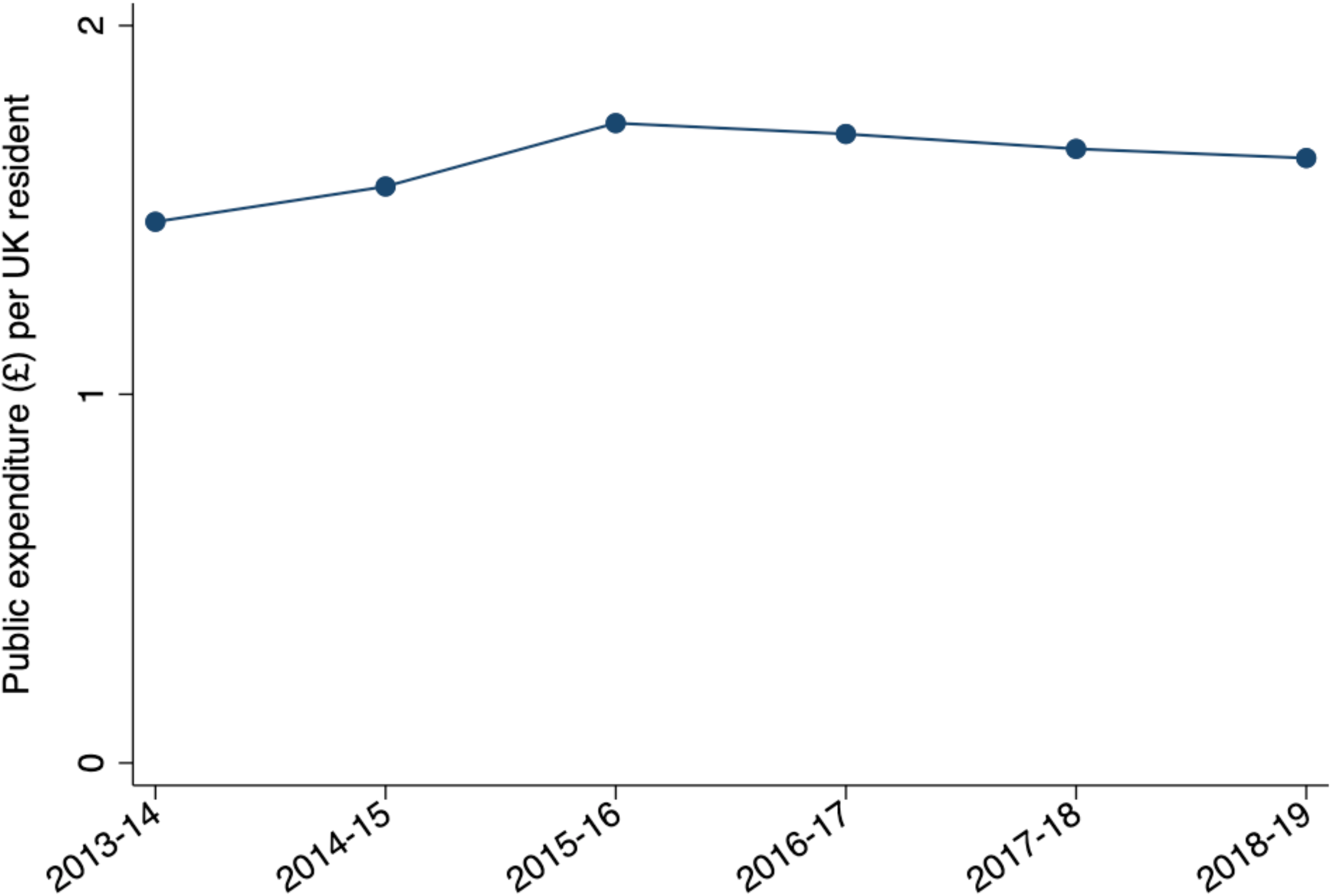
Public expenditure (£ per UK resident) on OTC codeine products in the UK between April 2013-March 2014 and April 2018-March 2019.

## DISCUSSION

Nearly 5 billion dosage units of codeine were sold over the counter (OTC) in the UK between April 2013 and March 2019. For every UK resident, the average per-person purchase was 694 mg of codeine tablets, 151 mL of codeine liquid, 26 g of paracetamol, and 1.4 g of ibuprofen over six years. During the study period, sales fell by one dosage unit per UK resident, but the cost to the public gradually increased. The UK market offered various of products containing codeine, tablet formulations being the most common. Three out of every four products contained paracetamol in combination with codeine. Over £600 million was spent on OTC codeine products in the UK between April 2013 and March 2019.

In this present study it was not possible to determine the specific indications for the billions of dosage units of codeine sold OTC; however, use was probably driven by several factors. Codeine is most often used to manage physical pain, including migraine, dental, back, menstrual, joint, postoperative, and childbirth pain [10,30,31]. However, codeine has also been reported to be used for sleep, relaxation, and reducing stress and depression [30,31], unlicensed indications. People who reported using codeine outside of its indications described becoming dependent on codeine because of environmental factors, including being unsupervised over a long time [10]. With OTC access to codeine, there is the risk of “pharmacy shopping” (i.e. purchasing single packets of codeine from multiple pharmacies), which has been reported in Ireland and the UK [30].

The growing pressures in the UK’s National Health Service (NHS), including long wait times and backlogs, have increased the need for self-care and self-medication [32–35]. Several public campaigns have been launched in the past decade to promote self-care, including most recently, the “Help Us Help You” campaign in February 2023, which was launched after finding that only one in five people aged between 18 and 40 accessed their local pharmacy for advice on minor illnesses [36]. While these initiatives are essential for the sustainability of the NHS, how such campaigns affect self-medication with products like codeine should be considered.

The slight fall in sales of codeine we found may be following the trends in prescribed opioids, which has also fallen [37,38]. However, the number of deaths attributed to codeine in England and Wales has increased in the last two decades [11], and the number of young people seeking help for codeine-related problems doubled between 2018 and 2019 in England [39]. Long-term users of codeine have reported that they were not aware of the risks and addictive potential of codeine when they first started taking it for pain [10]. Therefore, data on the sales and use of codeine should be monitored as the growing burden of pain and the need to self-medicate continue to increase.

A retrospective observational study of OTC codeine sales in 31 countries showed that the UK had the fourth highest sales volume [22]. Before this study, there was little data on the history of sales of OTC codeine in the UK. A report by the UK’s MHRA stated that an analysis of OTC codeine sales was to be conducted before and after regulatory changes in 2009 [19], but that analysis never took place. Although the MHRA made recommendations to reduce pack sizes of codeine to 32 tablets in 2005, we still identified sales that contained up to 100 tablets and liquid formulations up to 2 L, which has significant safety concerns.

In August 2023, the UK’s MHRA opened a public consultation on the availability of codeine linctus [18]. If the status of codeine changes from OTC to prescription-only in the UK, lessons from other countries that have made such changes, including France and Australia [14,17], should be considered, such as programmes to support people already dependent on codeine [40]. If the status of codeine does not change in the UK, the real-time monitoring of codeine sales in community pharmacies should identify safety concerns alongside the regular surveillance of adverse events reported from hospital admissions and the MHRA’s Yellow Card Scheme. Successful real-time monitoring programmes, including Australia’s ProjectSTOP for pseudoephedrine [41,42], could be used as a framework for monitoring codeine sales in the UK.

### Strengths and Limitations

To the best of our knowledge, this study provides the first in-depth analysis of OTC codeine sales in the UK. The data were obtained from a commercial company, *IQVIA*, which collects information via barcodes directly from providers. Owing to contractual agreements with *IQVIA*, the data cannot be openly shared. However, we preregistered our study protocol and openly shared the study materials via a repository [28]. The impact of the Covid-19 pandemic on the sales of OTC codeine cannot be ascertained from this study as the data were purchased in 2019. At the time of data provision, *IQVIA* reported that their coverage represented 67% of providers in the UK; however, this was shared at a single time and may have changed over time. Given the commercial nature of such data, *IQVIA* withheld methodological information about their data, including the formula used for calculating dosage units. Codeine can also be prescribed and purchased via the dark web or the internet [31], which was not captured in this dataset. The data provided by *IQVIA* were for products indicated for adults, but the annual population for all ages including children was used to calculate the sales rate over time. This analysis provides insights into sales and expenditure at the population level, which can only be a proxy for the actual use of OTC codeine in the community. Thus, it is not possible to determine the causal extent of codeine-related harms from the high volume of OTC codeine sales that we identified in the UK.

## CONCLUSIONS

This retrospective observational study illustrates that it is possible to capture data on the sales of OTC medicines in the UK. The MHRA and the government should consider the substantial quanitity of codeine sold in the UK as they examine the prescription status and availability. In the meantime, data on the sales of OTC codeine should be accessible and monitored to identify safety concerns.

## ACKNOWLEDGMENTS

We thank IQVIA for collecting and providing the data used in this study.

## AUTHOR CONTRIBUTIONS

GCR conceptualised, designed, and initiated the study, conducted all data analyses, created the figures, and edited the first draft of the manuscript. IEM wrote the first draft of the manuscript and revised subsequent drafts. JKA and CH provided supervisory support and oversight. All study authors read, contributed to, and approved the final manuscript.

## DATA AVAILABILITY STATEMENT

Data cannot be made openly available because of contractual agreements with IQVIA, but they can be accessed directly from IQVIA for a fee. All study materials are openly available on an open repository (https://doi.org/10.17605/OSF.IO/APG8E) [28].

## REFERENCES

[1] Jani M, Yimer BB, Sheppard T, Lunt M, Dixon WG. Time trends and prescribing patterns of opioid drugs in UK primary care patients with non-cancer pain: A retrospective cohort study. PLoS Med 2020;17. 10.1371/JOURNAL.PMED.1003270.

[2] Co-codamol for adults: painkiller containing paracetamol and codeine - NHS n.d. https://www.nhs.uk/medicines/co-codamol-adults/ (accessed July 25, 2023).

[3] Moore RA, Wiffen PJ, Derry S, Maguire T, Roy YM, Tyrrell L. Non-prescription (OTC) oral analgesics for acute pain - an overview of Cochrane reviews. Cochrane Database of Systematic Reviews 2015;2017. 10.1002/14651858.CD010794.PUB2.

[4] Derry S, Moore RA, McQuay HJ. Single dose oral codeine, as a single agent, for acute postoperative pain in adults. Cochrane Database of Systematic Reviews 2010;2019. 10.1002/14651858.CD008099.PUB2.

[5] Omar MI, Alexander CE. Drug treatment for faecal incontinence in adults. Cochrane Database of Systematic Reviews 2013;2013. 10.1002/14651858.CD002116.PUB2.

[6] Molassiotis A, Bailey C, Caress A, Tan JY. Interventions for cough in cancer. Cochrane Database of Systematic Reviews 2015;2015. 10.1002/14651858.CD007881.PUB3.

[7] Codeine phosphate. BNF n.d. https://bnf.nice.org.uk/drugs/codeine-phosphate/#indications-and-dose (accessed July 20, 2023).

[8] Codeine Addiction And Abuse - Addiction Center n.d. https://www.addictioncenter.com/opiates/codeine/ (accessed July 27, 2023).

[9] Agnich LE, Stogner JM, Miller BL, Marcum CD. Purple drank prevalence and characteristics of misusers of codeine cough syrup mixtures. Addictive Behaviors 2013;38:2445–9. 10.1016/J.ADDBEH.2013.03.020.

[10] Kinnaird E, Kimergård A, Jennings S, Drummond C, Deluca P. From pain treatment to opioid dependence: a qualitative study of the environmental influence on codeine use in UK adults. BMJ Open 2019;9. 10.1136/BMJOPEN-2018-025331.

[11] Codeine deaths England & Wales 2021. Statista 2021. https://www.statista.com/statistics/470902/death-by-codeine-drug-poisoning-in-england-and-wales/ (accessed July 20, 2023).

[12] Foley M, Harris R, Rich E, Rapca A, Bergin M, Norman I, et al. The availability of over-the-counter codeine medicines across the European Union. Public Health 2015;129:1465–70. 10.1016/J.PUHE.2015.06.014.

[13] Jouanjus E, Gibaja V, Fabre F, Lapeyre-Mestre M, Perri-Plandé J, Le Boisselier R, et al. Medical prescription forms of opioid cough suppressants falsified by the patients before and after they switched from over-the-counter to prescription-only in France. Br J Clin Pharmacol 2022;88:1713–21. 10.1111/BCP.15052.

[14] Pain S, Fauconneau B, Ricou J, Pérault-Pochat M-C. Changement de règlementation sur la codéine en 2017 en France : quelles conséquences pour les pharmaciens ? Therapies 2021;76:164. 10.1016/J.THERAP.2021.01.016.

[15] Jouanjus E, Gibaja V, Fabre F, Lapeyre-Mestre M, Perri-Plandé J, Le Boisselier R, et al. Medical prescription forms of opioid cough suppressants falsified by the patients before and after they switched from over-the-counter to prescription-only in France. Br J Clin Pharmacol 2022;88:1713–21. 10.1111/BCP.15052.

[16] Lenormand A. Les effets de la nouvelle règlementation de la codéine sur sa consommation n.d.

[17] Cairns R, Schaffer AL, Brown JA, Pearson SA, Buckley NA. Codeine use and harms in Australia: evaluating the effects of re-scheduling. Addiction 2020;115:451–9. 10.1111/ADD.14798.

[18] MHRA launches public consultation on reclassification of opioid-containing cough medicine - GOV.UK n.d. https://www.gov.uk/government/news/mhra-launches-public-consultation-on-reclassification-of-opioid-containing-cough-medicine (accessed July 25, 2023).

[19] MHRA. Codeine and dihydrocodeine-containing medicines: minimising the risk of addiction 2009. https://assets.publishing.service.gov.uk/media/5df7610c40f0b60956f5a2e4/Codeine_and_dihydrocodeine_minimising_the_risk_of_addiction.pdf (accessed August 9, 2023).

[20] Codeine: restricted use as analgesic in children and adolescents after European safety review - GOV.UK n.d. https://www.gov.uk/drug-safety-update/codeine-restricted-use-as-analgesic-in-children-and-adolescents-after-european-safety-review (accessed October 4, 2023).

[21] Opioid Expert Working Group meets at MHRA. Medicines and Healthcare Products Regulatory Agency n.d. https://www.gov.uk/government/news/opioid-expert-working-group-meets-at-mhra (accessed October 4, 2023).

[22] Richards GC, Aronson JK, MacKenna B, Goldacre B, Hobbs FDR, Heneghan C. Sales of Over-the-Counter Products Containing Codeine in 31 Countries, 2013–2019: A Retrospective Observational Study. Drug Saf 2022;45:237–47. 10.1007/S40264-021-01143-2/

[23] Richards GC, Goldacre B, Curtis H, Hobbs R, Perera-Salazar R, Aronson JK, et al. Over-the-counter (OTC) codeine & the UK opioid crisis 2019. 10.17605/OSF.IO/D6TBH

[24] Gery P. Guy Jr, Haegerich TM, Evans ME, Losby JL, Young R, Jones CM. Vital Signs: Pharmacy-Based Naloxone Dispensing — United States, 2012–2018. Morbidity and Mortality Weekly Report 2019;68:679. 10.15585/MMWR.MM6831E1.

[25] McKee KA, Crocker CE, Tibbo PG. Long-acting injectable antipsychotic (LAI) prescribing trends during COVID-19 restrictions in Canada: a retrospective observational study. BMC Psychiatry 2021;21:1–10. 10.1186/S12888-021-03646-9/FIGURES/3.

[26] Olfson M, Waidmann T, King M, Pancini V, Schoenbaum M. Population-Based Opioid Prescribing and Overdose Deaths in the USA: an Observational Study. J Gen Intern Med 2023;38:390–8. 10.1007/S11606-022-07686-Z/TABLES/4.

[27] Jayawardana S, Forman R, Johnston-Webber C, Campbell A, Berterame S, de Joncheere C, et al. Global consumption of prescription opioid analgesics between 2009-2019: a country-level observational study. EClinicalMedicine 2021;42:101198. 10.1016/j.eclinm.2021.101198.

[28] Richards C G, Curtis H, Aronson K J, Heneghan C, Martus I. OTC codeine sales in the UK. OSF 2023. 10.17605/OSF.IO/APG8E (accessed August 11, 2023).

[29] Richards GC. Sales of over-the-counter (OTC) codeine-containing products in 31 countries: georgiarichards/otc_codeine. Github 2021. 10.5281/ZENODO.5720700.

[30] Kimergard A, Foley M, Davey Z, Dunne J, Drummond C, Deluca P. Codeine use, dependence and help-seeking behaviour in the UK and Ireland: an online cross-sectional survey. QJM: An International Journal of Medicine 2017;110:559–64. 10.1093/QJMED/HCX076.

[31] Van Hout MC, Hearne E. Confessions of contemporary English opium-eaters: a netnographic study of consumer negotiation of over-the-counter morphine for misuse. J Subst Use 2016;21:141–52. 10.3109/14659891.2014.980861.

[32] NHS waiting times: our position. The King’s Fund 2022. https://www.kingsfund.org.uk/projects/positions/nhs-waiting-times (accessed October 10, 2023).

[33] Self-help guide: Lower back pain | NHS inform n.d. https://www.nhsinform.scot/self-help-guides/self-help-guide-lower-back-pain (accessed August 6, 2023).

[34] NHS backlog data analysis. British Medical Association n.d. https://www.bma.org.uk/advice-and-support/nhs-delivery-and-workforce/pressures/nhs-backlog-data-analysis (accessed October 10, 2023).

[35] NHS England. Encouraging people to choose self care for life 2018. https://www.england.nhs.uk/2018/11/encouraging-people-to-choose-self-care-for-life/ (accessed October 4, 2023).

[36] NHS. NHS launches ad campaign as just one in five would visit high-street pharmacy for minor illnesses. NHS England 2023. https://www.england.nhs.uk/2023/02/nhs-launches-ad-campaign-as-just-one-in-five-would-visit-high-street-pharmacy-for-minor-illnesses/ (accessed October 13, 2023).

[37] Curtis HJ, Croker R, Walker AJ, Richards GC, Quinlan J, Goldacre B. Opioid prescribing trends and geographical variation in England, 1998–2018: a retrospective database study. Lancet Psychiatry 2019;6:140–50. 10.1016/S2215-0366(18)30471-1.

[38] Martus I;, Mackenna B;, Rial W;, Hayhurst J;, Richards G. Private prescribing of controlled opioids in England. BJGP 2023:2014–21. 10.3399/BJGP.2023.0146.

[39] GOV.UK. Young people’s substance misuse treatment statistics 2020 to 2021: report. Office of Health Improvement & Disparities 2022. https://www.gov.uk/government/statistics/substance-misuse-treatment-for-young-people-statistics-2020-to-2021/young-peoples-substance-misuse-treatment-statistics-2020-to-2021-report (accessed October 19, 2023).

[40] Supporting patients with codeine dependence. NPS MedicineWise 2020. https://www.nps.org.au/professionals/codeine/supporting-patients-with-codeine-dependence (accessed July 25, 2023).

[41] Hattingh HL, Varsani J, Kachouei LA, Parsons R. Evaluation of pseudoephedrine pharmacy sales before and after mandatory recording requirements in Western Australia: A case study. Subst Abuse Treat Prev Policy 2016;11. 10.1186/s13011-016-0075-0.

[42] Ferris J, Devaney M, Mazerolle L, Sparkes-Carroll M. Assessing the utility of Project STOP in reducing pseudoephedrine diversion to clandestine laboratories. Australian Institue of Criminology 2016. https://www.aic.gov.au/publications/tandi/tandi509.

